# Impact of Aspirin Therapy on Progression of Thoracic and Abdominal Aortic Aneurysms

**DOI:** 10.1101/2025.02.05.25321726

**Authors:** Shayan Mohammadmoradi, Kory Heier, Elizabeth R. Driehaus, Hammodah R. Alfar, Sam Tyagi, Kristen McQuerry, Sidney W. Whiteheart

## Abstract

**Background and Objective:** Aortic aneurysms, including abdominal (AAA) and thoracic (TAA), pose significant challenges due to their rupture risk and complex pathophysiology. While aspirin has been proposed to manage aneurysm progression, evidence remains limited. This retrospective, single-center study used AI-driven methods to examine the association between aspirin therapy and aneurysm growth.

**Methods:** The study, at the University of Kentucky Healthcare, utilized de-identified electronic health record data from 2010 to 2023. To evaluate platelet counts changes, Cohort 1 included patients with AAA or TAA and matched healthy controls. Cohort 2 included AAA or TAA patients who had at least two imaging studies. Extraction of aortic diameters utilized an advanced AI-based natural language processing (NLP) algorithm to identify and extract relevant text strings related to aortic dimensions. Multivariable-adjusted linear regression analyses assessed the impact of aspirin on aneurysm progression.

**Results:** Cohort 1 included 11,538 participants: 5,774 controls, 3,439 with AAA, and 2,325 with TAA. Platelet counts were significantly lower in patients with aortic aneurysms compared to controls, though they were not considered thrombocytopenic. Cohort 2 included 302 AAA and 141 TAA patients. Subgroup analysis revealed that aspirin use was associated with increased AAA progression in females with small aneurysms (<50 mm). Further, aspirin therapy showed no significant impact on the annualized change in aneurysm diameter for TAA or for males with AAA.

**Conclusion:** Our findings suggest aspirin’s effectiveness varies by sex and potentially aneurysm size, underscoring the need for further research to refine antiplatelet therapy guidelines for aortic aneurysms.

## Introduction

Aortic aneurysms represent a significant challenge in cardiovascular medicine, given their complex pathophysiology and the potentially catastrophic consequences of rupture. ^1–3^ Current management focuses on early diagnosis, surveillance, and surgical repair, while pharmacological therapies to slow aneurysm growth remain underdeveloped. ^4–6^ Platelet activation and the coagulation cascade play critical roles in aneurysm progression. ^2,7–10^ Given their role in vascular homeostasis, platelets and coagulation factors may play critical but less studied roles in aortic aneurysm progression.

In abdominal aortic aneurysms (AAA), platelets contribute to intraluminal thrombus (ILT) formation and release inflammatory mediators and proteolytic enzymes that degrade the extracellular matrix and weaken the aortic wall. ^2,8,9,11^ Thoracic aortic aneurysms (TAA), although involving minimal ILT, exhibit significant platelet activation, which directly impairs endothelial integrity and amplifies inflammation.^12^ Antiplatelet therapies, traditionally used for their antithrombotic effects, may also target these pathological pathways, potentially stabilizing aneurysm growth. ^2,13,14^ The 2022 American College of Cardiology and American Heart Association guidelines recommend aspirin with a class 2b indication (level of evidence: C) for patients with AAA and intramural thrombus. ^5^ Similarly, the European Society for Vascular Surgery advises the use of antiplatelet agents in patients undergoing AAA surgery, though evidence supporting their impact on sac volume remains limited. ^6^

Studies of antiplatelet therapies have focused on AAA, with comparatively less emphasis on TAA, and they often lack clarity on sex differences. ^3,8,9,15,16^ Such studies are also hindered by challenges in data collection and analysis. Advances in artificial intelligence, particularly natural language processing (NLP) ^17–20^, offer new opportunities to leverage electronic health records (EHRs) for studying aneurysm progression and therapeutic efficacy. NLP—a branch of artificial intelligence—has gained traction for transforming unstructured data, such as imaging reports, into analyzable variables. ^18^ It is particularly valuable in aneurysm research, where imaging reports often detail aortic diameters that can be correlated with treatment history and disease progression.

To address these gaps in understanding the effectiveness of anti-platelet therapies, our retrospective single-center study utilizes a novel AI-driven data extraction method to obtain aortic diameter measurements. By analyzing the association between aspirin therapy and aneurysm growth rates in both AAA and TAA patients, with a focus on sex differences, we provide more insights to the therapeutic potential of antiplatelet agents in managing aortic aneurysms.

## Methods

The data supporting the findings of this study are available from the corresponding author upon reasonable request.

### Study Design and Population

This study adheres to the Strengthening the Reporting of Observational Studies in Epidemiology (STROBE) guidelines for cohort studies, ensuring comprehensive reporting. Conducted as a retrospective cohort analysis at the University of Kentucky Healthcare (UK Healthcare), the study utilized de-identified data extracted from the electronic health records of patients diagnosed with AAA and TAA. The data were extracted from electronic health records using *International Statistical Classification of Disease and Related Health Problems, Tenth Revision* (ICD-10) codes for aortic aneurysms (**Supplementary Table 1**). The data collection spanned 2010 to 2023 and was facilitated through the UK Center for Clinical and Translational Science (CCTS). The University of Kentucky Institutional Review Board approved this study and waived the need for patient consent due to its retrospective design. All patient information was de-identified prior to analysis and reported in aggregate.

The Cohort 2 study population comprised patients with diagnoses of AAA or TAA (based on the ICD-10 codes) without rupture who had undergone at least two imaging screens to monitor aneurysm size over time. Exclusion criteria included patients with prior aneurysm repair, and those with incomplete health records. A summary of the selection criteria for the final analysis population is presented in **Figure 1**. Patient race, documented as self-reported in their electronic health records, was categorized as Black, White, and Other (including American Indian, Asian, Hispanic, or Pacific Islander).

**Figure 1:**
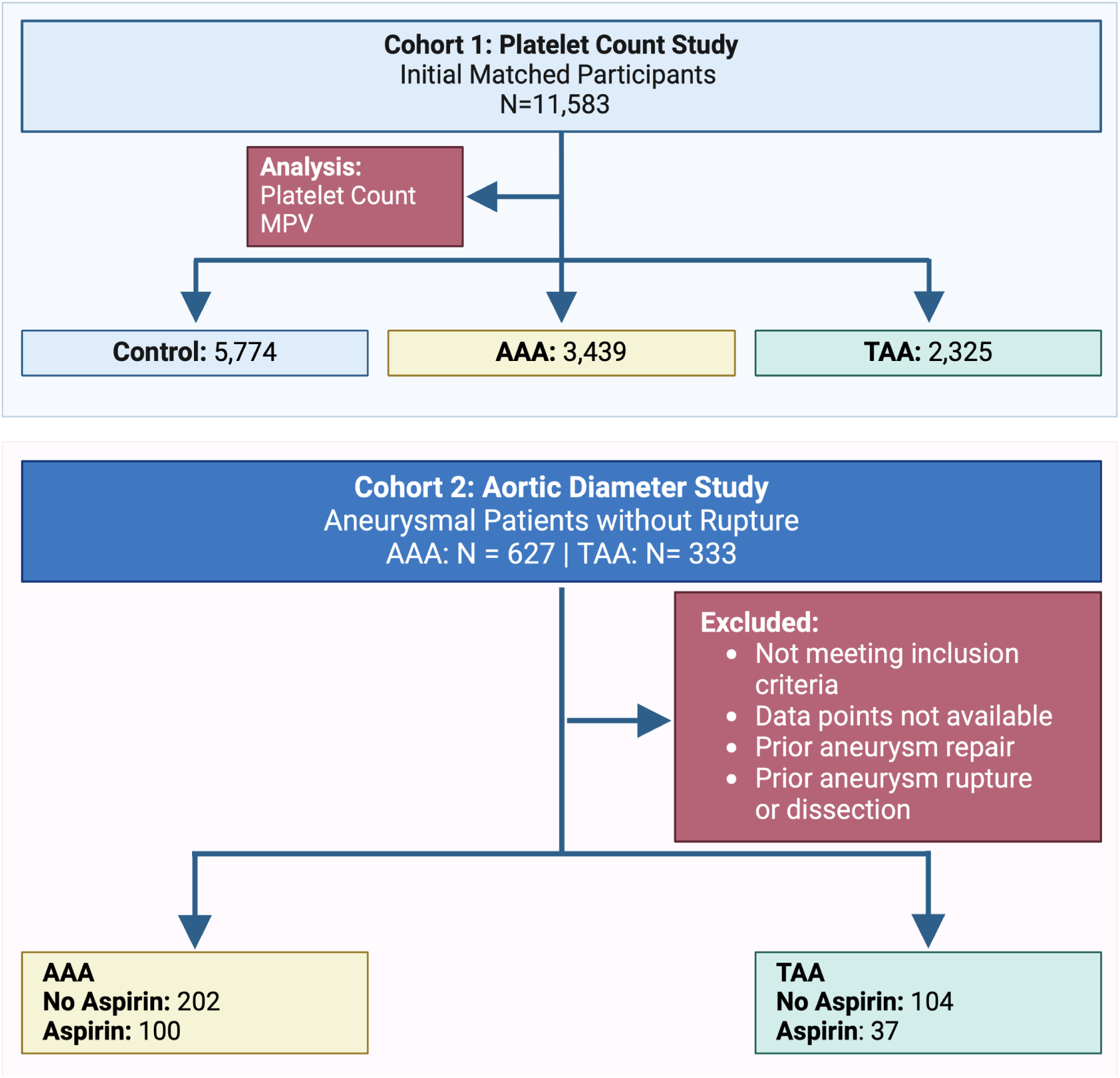
Flowchart of the two study cohorts. **Cohort 1** (platelet count study) consisted of patients and their matched controls, with available demographic data, platelet counts, and mean platelet volume (MPV) measurements. **Cohort 2** (aortic diameter study) included patients with either AAA or TAA who received aspirin therapy and had at least two imaging assessments available from the University of Kentucky Healthcare database.

### AI-based Methodology for Extracting Aortic Diameter Measurements

A dataset of aortic diameters was created by extracting measurements from medical reports of patients who underwent imaging at the University of Kentucky HealthCare Radiology department. Patients received imaging using either ultrasound or computed tomography (CT) scans, following the institution’s standard operating procedures. All imaging studies were reviewed and overread by a registered physician specialized in vascular interpretation to ensure accuracy and reproducibility of diameter measurements, per institutional standard. The dataset consisted of anonymized medical imaging reports from patients with diagnosed aortic aneurysms, and textual narratives of the imaging findings and impressions. Preliminary data processing and analysis were performed using Python. The extraction algorithm was built using OpenAI’s GPT-4 (Generative Pretrained Transformer) technology, leveraging its advanced capabilities in text understanding and pattern recognition (OpenAI, 2024, “Generative Pretrained Transformer Technology for Natural Language Processing,” URL: chat.openai.com). This sophisticated natural language processing (NLP) model enabled precise identification of relevant text within the narrative sections of imaging reports, focusing on keywords and numerical expressions related to aortic dimensions. The code snippets used for data extraction are provided in the supplementary material.

A curated list of keywords associated with aortic aneurysms and related conditions (e.g., “ascending thoracic aorta,” “abdominal aortic aneurysm”) was compiled based on clinical relevance. Regular expression (regex) techniques were employed to detect and extract numerical values denoting the aortic diameter, immediately following these keywords within the text. The regex patterns were designed to capture both the measurement (in mm or cm) and the associated unit, allowing for the conversion of all values to millimeters for consistency. To enhance the specificity of the extraction, the algorithm was programmed to ignore measurements associated with non-target anatomical features or pathologies (e.g., “liver size,” “spleen length”).

The extraction process was implemented in Python on the ChatGPT-4 platform, utilizing its string manipulation capabilities to parse and analyze the text data. The algorithm iteratively processed each report, applying the regex-based extraction to the patient report text. The maximum value identified was recorded as the maximal aortic diameter measurement for each patient encounter. In instances where no relevant aortic diameter measurement was identified, the record was annotated with “ND” (Not Detected). This step ensured that the dataset comprehensively represented both the presence and absence of quantifiable aortic measurements, facilitating subsequent analyses. To the best of our knowledge, this study represents the first application of OpenAI’s ChatGPT in extracting aortic diameter measurements from deidentified medical imaging reports. The high accuracy rate underscores the potential of advanced NLP models to significantly streamline the analysis of medical text data.

### Data Management

Two cohorts of data were collected from the University of Kentucky Center for Clinical and Translational Science (CCTS) Database. Each dataset included demographic information, clinical characteristics, comorbidities, medication usage, and imaging reports. Cohort 1 (platelet count study) contained control, TAA, and AAA patients with their corresponding platelet count lab values. Cohort 2 (aortic diameter study) contained patient information about aortic diameters of patients who underwent imaging at least twice at University of Kentucky Healthcare. The extraction process utilized an advanced AI-based natural language processing (NLP) algorithm to identify and extract relevant text strings related to aortic dimensions from medical imaging reports. The accuracy of the automated extraction process was validated through manual review, achieving a 98% concordance rate.

### Statistical Analyses

All data were analyzed using SAS v9.4 (SAS, Cary, NC, USA) and R programming language, version 4.1.1 (R Foundation for Statistical Computing, Vienna, Austria). All statistical tests were two-sided and statistical significance was defined as p-value ≤0.05. Progression was defined as the change in mean, annualized diameter.

Demographic and clinical characteristics were analyzed based on subject’s earliest admission date. Subjects included were those with a AAA and/or TAA diagnosis with at least 2 vascular images. The demographic table includes summaries for age, sex, race, baseline aneurysm diameter, comorbidities, and lab values separately for AAA and TAA stratified by aspirin use. Continuous variables were analyzed using Wilcoxon rank sum test and categorical variables were analyzed using a Chi-Square or Fisher’s exact test. Subjects who had first and last diameter measurements within 24 hours were excluded, and those subjects who had large negative change values without surgical intervention.

For each subject, the earliest order of lab performed date was analyzed. One-to-one propensity score matching was used to balance observed baseline covariates between control and aneurysm (first diagnosis of AAA with no rupture or TAA with no rupture) cohorts. A greedy algorithm was employed with a caliper of 0.25. Baseline covariates considered were age at first encounter, sex, race, COPD, type II diabetes, hypertensive, smoking status, and acute kidney failure. Data were analyzed using an ANOVA model for this subset of subjects with a Tukey adjustment for multiple comparisons.

Differences from first to last aneurysm diameter by drug therapy use were analyzed using the mean annualized change in aortic diameter using multivariable-adjusted linear regression analyses for drug therapy users. Covariates consist of age, sex, race, baseline aneurysm diameter, and comorbidities. These analyses were further stratified by sex and baseline diameter. The change in mean annualized diameter was derived by taking difference in the first and last measured diameter on the aneurysm imaging divided by the duration between measurements. Additionally, we performed a multivariable logistic regression to look at the association of drug therapy use with rapid progression, defined as an increase in diameter of 5 mm or more per year.

## Results

### Platelet Count Is Significantly Lower in Patients with Aortic Aneurysms

Alterations in platelet count can be indications of their consumption. To investigate differences in platelet counts among patients with aortic aneurysms, platelet count data from individuals undergoing routine medical examinations without reported abnormalities (ICD code Z00.00 – healthy controls) were compared with that from patients diagnosed with abdominal aortic aneurysms (AAA) and thoracic aortic aneurysms (TAA) without rupture **(Figure 1)**. A cohort of 11,538 participants (cohort 1) was identified using ICD-10 codes to assess platelet count **(Table 1)**. This matched cohort included 5,774 controls, 3,439 individuals with AAA, and 2,325 with TAA. Gender distribution was consistent across groups, with males comprising 69% of both the AAA and control groups and 67% of the TAA group. Demographic and clinical characteristics—such as age, gender, race, smoking status, hypertension, diabetes, and COPD—were included as covariates in the statistical analyses. The racial composition was predominantly White in all groups, accounting for 93% of the AAA group, 91% of the TAA group, and 92% of controls. Smoking prevalence was highest in the AAA group (10%) compared to 8.3% in controls and 5.3% in the TAA group. Hypertension prevalence was similar between the AAA (45%) and control (43%) groups but slightly lower in the TAA group (43%).

**Table 1.**
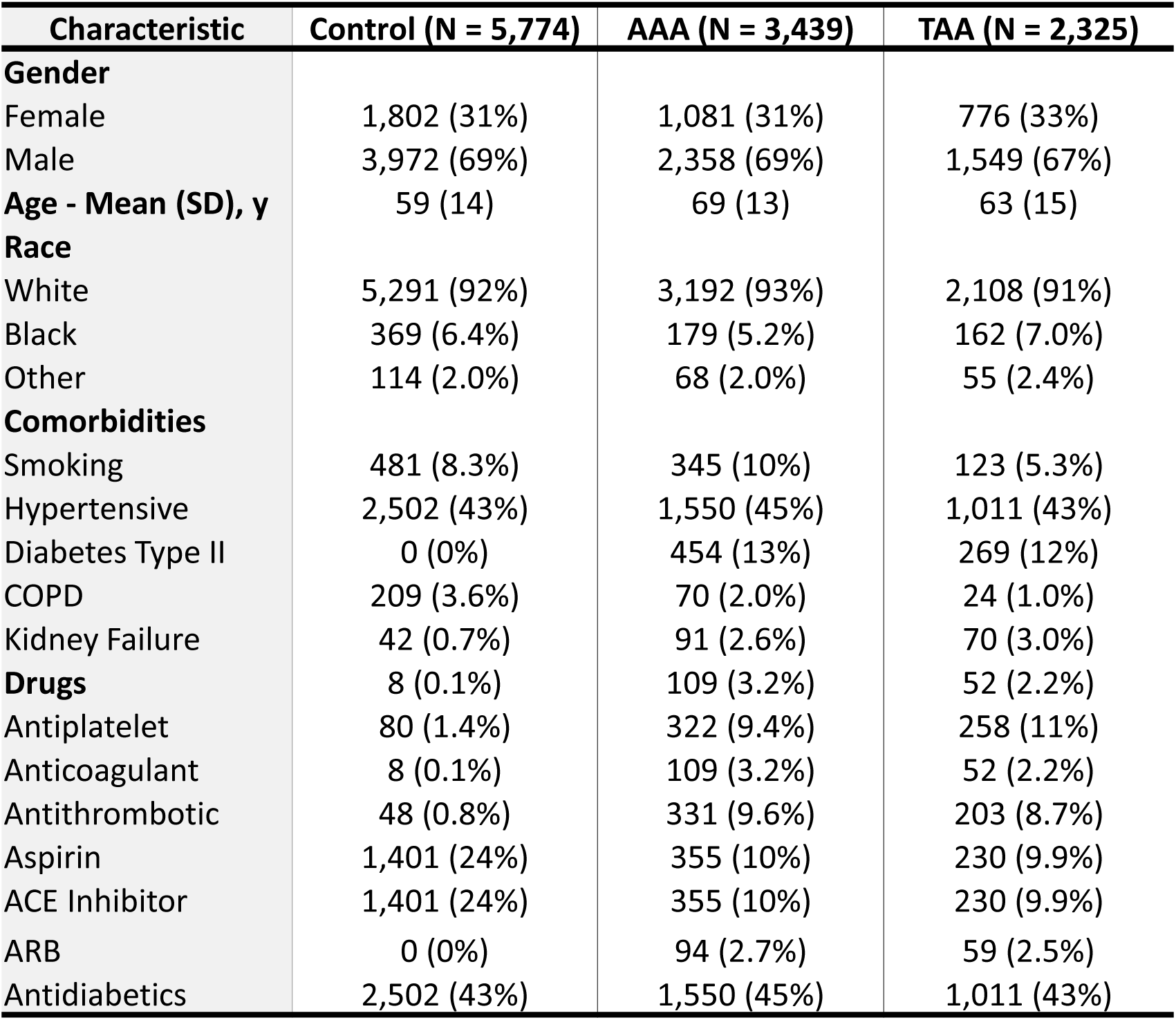
Demographic and clinical characteristics of the matched controls and aneurysm cohorts.

In cohort 1, data were analyzed using an ANOVA model. Comparisons, with a Tukey adjustment for multiple comparisons, between the control and AAA groups (Control: 236.27 ± 1.61 SE; AAA: 200.86 ± 1.78 SE), as well as between the control and TAA groups (Control: 236.27 ± 1.61 SE; TAA: 199.65 ± 2.15 SE), showed statistically significant lower levels of platelet counts (while still within normal range) in the aneurysmal groups (p < 0.001 for both). The difference between the AAA and TAA groups was not statistically significant (p = 0.90). Control subjects exhibited a mean platelet count of 247 ± 75 k/uL. In comparison, individuals with AAA and TAA had notably lower platelet counts, averaging 219 ± 95 k/uL and 217 ± 97 k/uL, respectively **(Figure 2-A)**. Furthermore, the mean platelet volume (MPV) showed minimal differences between the groups, with controls exhibiting an MPV of 10.48 ± 0.90 fL, AAA patients at 10.21 ± 0.95 fL, and TAA patients at 10.23 ± 0.99 fL **(Figure 2-B)**. It is important to note that while the reduction in platelet count is significant, these patients would not be considered clinically thrombocytopenic.

**Figure 2:**
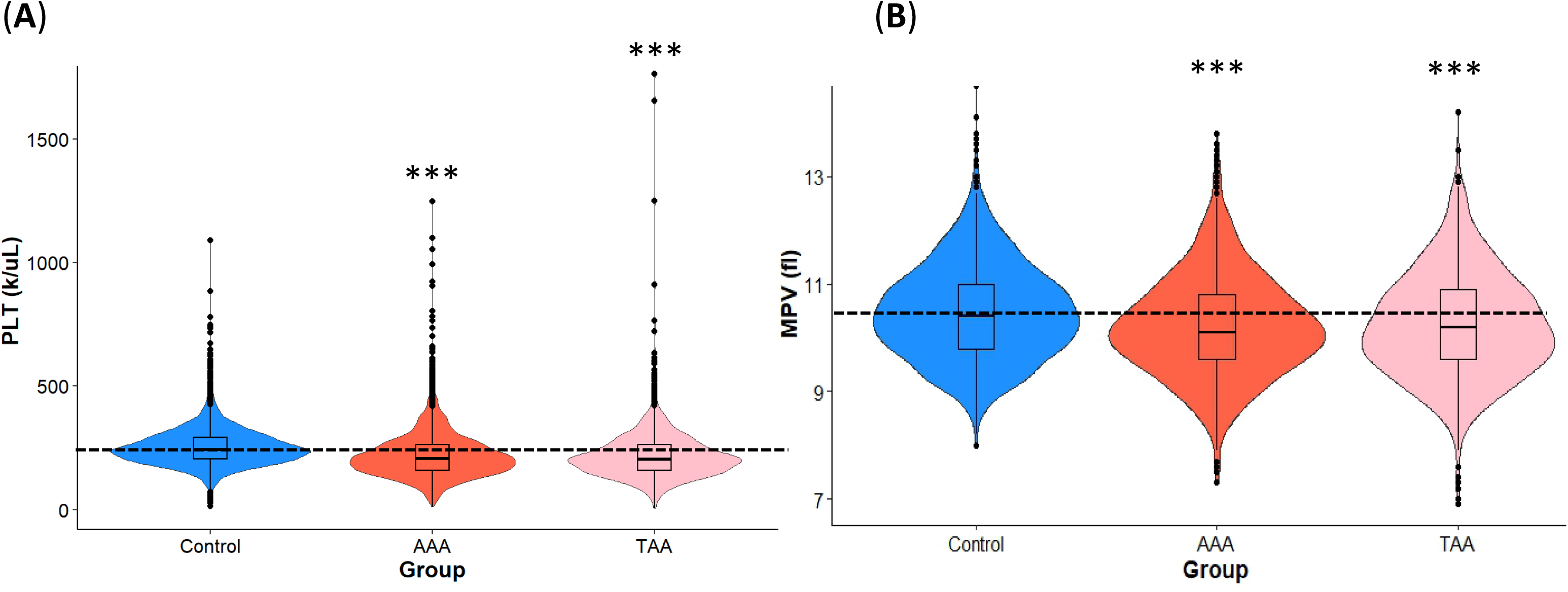
Platelet Counts and Mean Platelet Volumes Values Are Changed in Aneurysmal Patients without Rupture. Violin plots showing the distribution of platelet counts (PLT) **(A)** and Mean Platelet Volume (MPV) **(B)** among matched Control, AAA, and TAA groups. Embedded within each violin plot are boxplots that display the median (horizontal line inside the box), interquartile range (IQR, represented by the height of the box), and the points represent 1.5 times the IQR from the quartiles. (***) P<0.001 for control vs. AAA and control vs. TAA. Dotted line represents the median of Control group.

### Aspirin Affects AAA Progression

In cohort 2, Analysis of the impact of aspirin therapy on AAA and TAA progression involved patients with diagnoses based on their ICD-10 codes and available imaging reports from the University of Kentucky Healthcare database. Patients were stratified by their aspirin use, and differences in aneurysm progression were assessed using multivariable-adjusted linear regression analysis. We retrospectively analyzed 302 patients with AAA and 141 patients with TAA. Among the AAA cohort **(Table 2)**, 100 patients (32.2%) were on aspirin therapy, while in the TAA cohort (**Table 3**), 37 patients (25.0%) were receiving aspirin therapy. Baseline demographic and clinical characteristics were summarized across the two groups (**Table 2**, **3**).

**Table 2.** Demographic and clinical characteristics of the AAA cohort stratified by aspirin use.

| Characteristic | No Aspirin (N=202) | Aspirin (N=100) | p-value |
| --- | --- | --- | --- |
| **Gender** |  |  | 0.451 |
| Female | 62 (30.7%) | 35 (35.0%) |  |
| Male | 140 (69.3%) | 65 (65.0%) |  |
| **Age - Mean (SD), y** | 66.9 (10.5) | 66.6 (9.8) | 0.787 |
| **Race** |  |  | 0.606 |
| White | 182 (90.1%) | 94 (94.0%) |  |
| Black | 16 (7.9%) | 5 (5.0%) |  |
| Other | 4 (2.0%) | 1 (1.0%) |  |
| **Comorbidities** |  |  |  |
| Smoking | 45 (22.3%) | 31 (31.0%) | 0.1 |
| Hypertensive | 142 (70.3%) | 45 (45.0%) | **<0.001** |
| Diabetes Type II | 17 (8.4%) | 9 (9.0%) | 0.865 |
| COPD | 27 (13.4%) | 29 (29.0%) | **<0.001** |
| Acute Kidney Failure | 18 (8.9%) | 14 (14.0%) | 0.176 |
| **Baseline Diameter (mm)** |  |  |  |
| < 30 | 18 (8.9%) | 11 (11.0%) |  |
| 30 – 40 | 88 (43.6%) | 37 (37.0%) |  |
| 40 – 50 | 82 (40.6%) | 32 (32.0%) |  |
| > 50 | 14 (6.9%) | 20 (20.0%) |  |
| **AAA Rapid Growth** | 21 (10.4%) | 15 (15.0%) | 0.245 |
| **Platelet Count - Mean (SD)** | 221.9 (98.8) | 221.5 (79.3) | 0.973 |

**Table 3.**
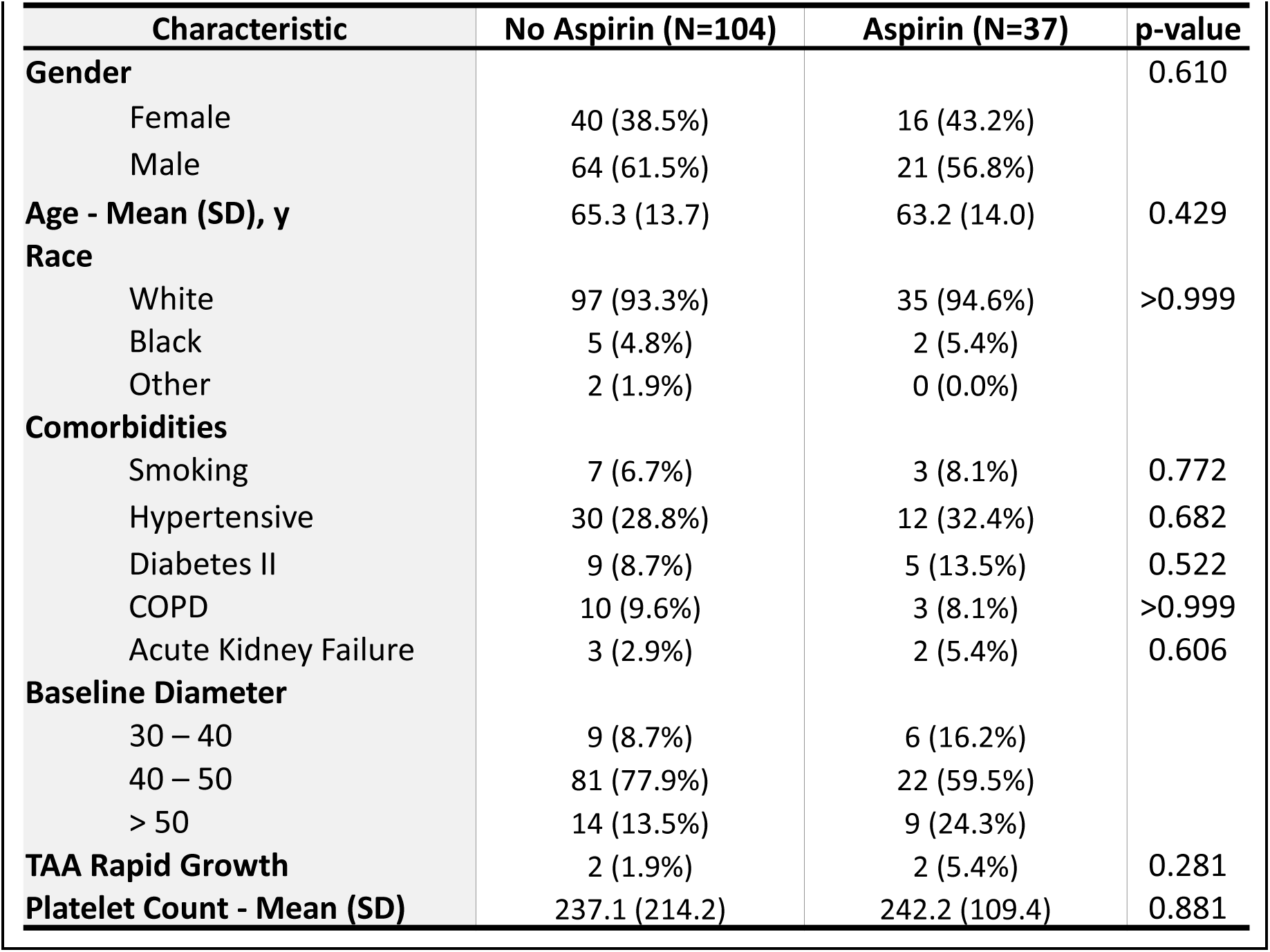
Demographic and clinical characteristics of the TAA cohort stratified by aspirin use.

| Characteristic | No Aspirin (N=104) | Aspirin (N=37) | p-value |
| --- | --- | --- | --- |
| **Gender** |  |  | 0.610 |
| Female | 40 (38.5%) | 16 (43.2%) |  |
| Male | 64 (61.5%) | 21 (56.8%) |  |
| **Age - Mean (SD), y** | 65.3 (13.7) | 63.2 (14.0) | 0.429 |
| **Race** |  |  |  |
| White | 97 (93.3%) | 35 (94.6%) | >0.999 |
| Black | 5 (4.8%) | 2 (5.4%) |  |
| Other | 2 (1.9%) | 0 (0.0%) |  |
| **Comorbidities** |  |  |  |
| Smoking | 7 (6.7%) | 3 (8.1%) | 0.772 |
| Hypertensive | 30 (28.8%) | 12 (32.4%) | 0.682 |
| Diabetes II | 9 (8.7%) | 5 (13.5%) | 0.522 |
| COPD | 10 (9.6%) | 3 (8.1%) | >0.999 |
| Acute Kidney Failure | 3 (2.9%) | 2 (5.4%) | 0.606 |
| **Baseline Diameter** |  |  |  |
| 30 – 40 | 9 (8.7%) | 6 (16.2%) |  |
| 40 – 50 | 81 (77.9%) | 22 (59.5%) |  |
| > 50 | 14 (13.5%) | 9 (24.3%) |  |
| **TAA Rapid Growth** | 2 (1.9%) | 2 (5.4%) | 0.281 |
| **Platelet Count - Mean (SD)** | 237.1 (214.2) | 242.2 (109.4) | 0.881 |

In AAA patients, the mean age was 66.9 years in the non-aspirin group and 66.6 years in the aspirin group, with no significant differences between the groups (p = 0.787). Gender distribution was similar, with 69.3% male in the non-aspirin group and 65.0% male in the aspirin group (p = 0.451). Most patients were White (90.1% in the non-aspirin group and 94.0% in the aspirin group), with no significant differences in racial distribution (p = 0.606). Key comorbidities included hypertension (29.7% in the non-aspirin group and 55.0% in the aspirin group, p < 0.001), diabetes mellitus type II (8.4% in the non-aspirin group and 9.0% in the aspirin group, p = 0.865), and COPD (13.4% in the non-aspirin group and 29.0% in the aspirin group, p < 0.001).

Multivariable-adjusted linear regression analyses revealed a significant difference in the mean annualized change in diameter between aspirin users and non-users in the AAA cohort, but not in the TAA cohort (**Figure 3**). In the AAA cohort, the mean annualized change in diameter was significantly higher for aspirin users (3.55 mm/year ± 0.91 SE) compared to non-users (2.42 mm/year ± 0.85 SE; p = 0.031). However, in the TAA cohort, there was no significant difference between the groups (No Aspirin: 0.48 mm ± 0.70 SE, Aspirin: 1.14 mm ± 0.75 SE; p = 0.077). The least squares means for each level of aspirin administration indicate that aspirin use is associated with accelerated AAA growth, but not TAA growth.

**Figure 3:**
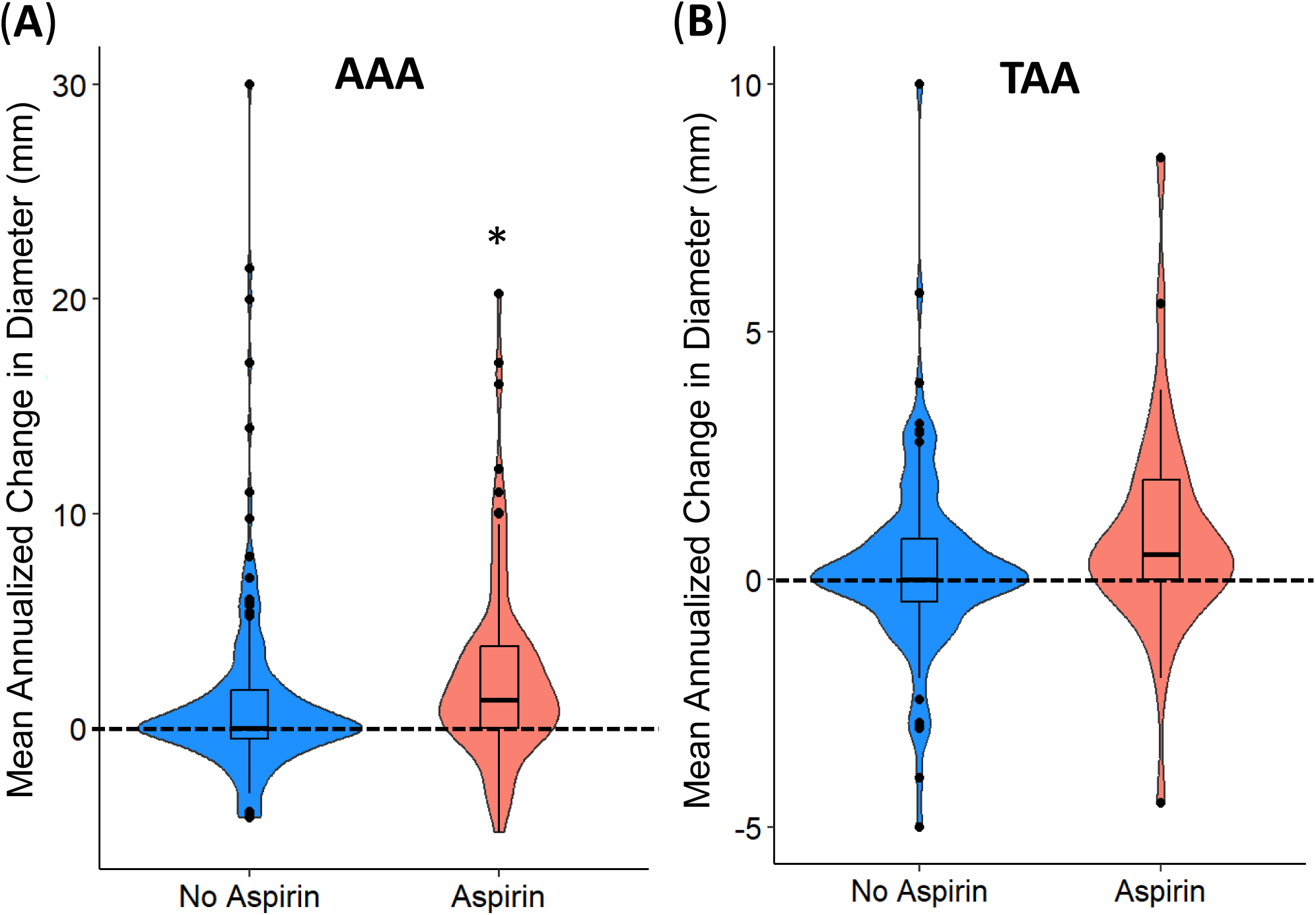
Aspirin Therapy in AAA Patients Resulted in Significant Alterations Annualized Aortic Diameter. Violin Plots of Mean Annualized Change in AAA (**A**) and TAA (**B**) Diameter by group utilizing a multivariable-adjusted linear regression analyses. Presented in the table are the least squares means for each group. (*) P < 0.05 was considered significant. Dotted line represents the median of Control group.

Our study further assessed the incidence of rapid progression, defined as an increase in aortic diameter of ≥ 5 mm per year. Neither AAA nor TAA aspirin users showed a significant difference in the odds of rapid progression compared to non-users. For AAA, the odds ratio (OR) was 1.07 [95% CI: 0.46, 2.39] (p = 0.873), and for TAA, the OR was 4.78 [95% CI: 0.34, 88.3] (p = 0.231).

Interestingly, subgroup analysis revealed that females receiving aspirin responded differently compared to males (**Figure 4**). Female participants on aspirin therapy showed a significant increase in the rate of AAA progression, with an average increase of 2.64 mm per year compared to non-aspirin users (p = 0.003). This effect was not seen in the male subgroup, where changes were not statistically significant.

**Figure 4:**
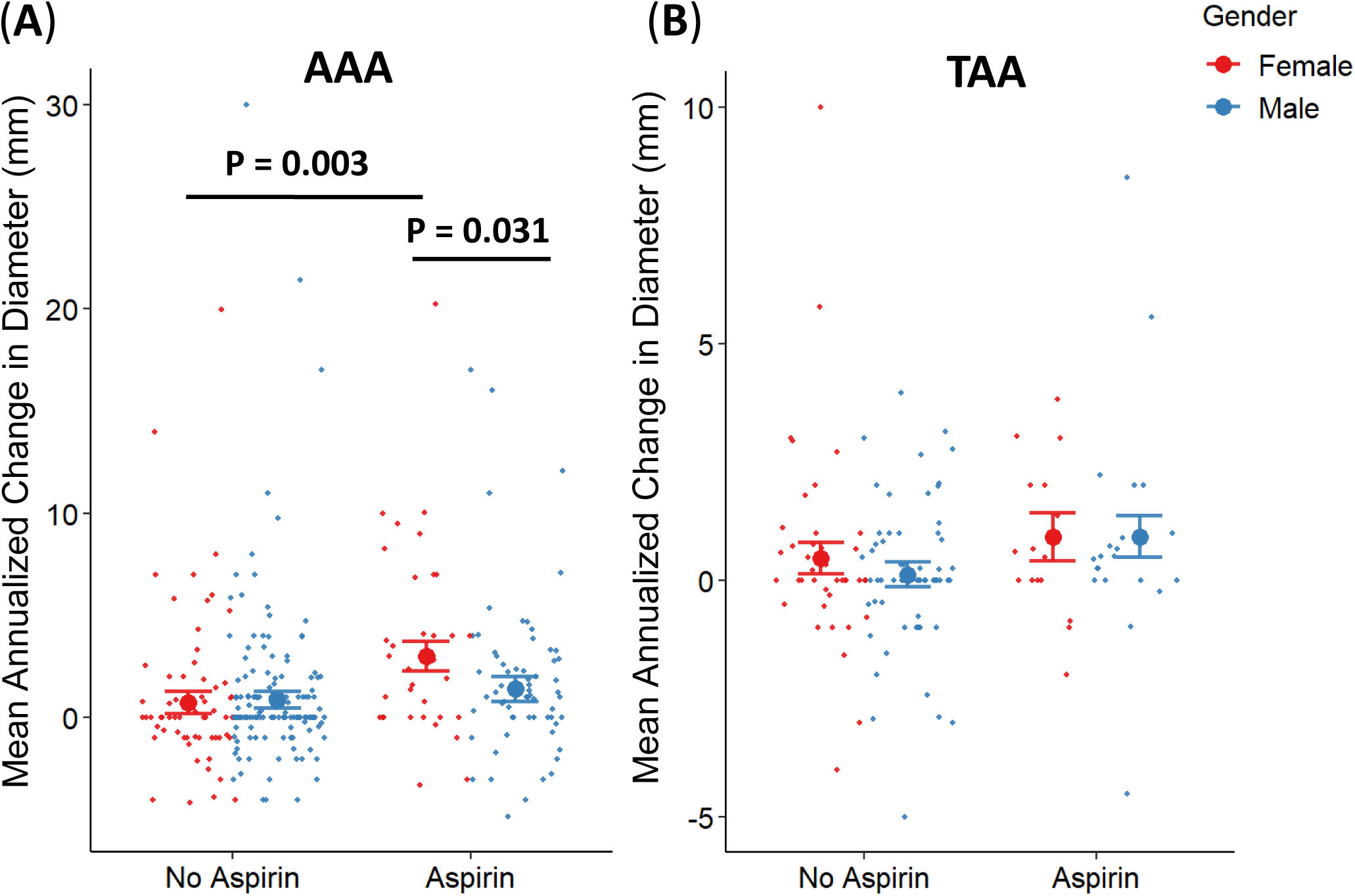
Annualized Growth for Aspirin Treatment and Gender Effect. Regression analysis of the distribution of annualized growth rates of AAA (**A**) and TAA (**B**) among patients, stratified by aspirin use and gender.

## Discussion

Our study investigated the potential role of platelets in the progression of both AAA and TAA, while also highlighting a previously unreported influence of sex on aneurysm progression outcomes following aspirin therapy. Few studies have explored the association between aspirin use and clinical outcomes in patients with aortic aneurysms, and those that do are often limited by heterogeneity in aneurysm size, a predominant focus on AAA rather than TAA, or a lack of attention to sex-specific differences in therapeutic outcomes. ^13–15,21,22^

Our findings revealed significant differences in platelet counts between control subjects and patients with aortic aneurysms, aligning with prior research that emphasizes the role of platelet activation in aneurysmal pathology.^1,7,15,23^ It should be noted that decrease in platelet count was no sufficient to consider patients thrombocytopenic. The majority of aneurysmal patients in our cohort had small aneurysms (<50 mm), allowing us to discern that while platelet activation may play a crucial role in aneurysm progression, the effectiveness of antiplatelet therapy—such as aspirin—varies based on aneurysm size and patient sex. These results underscore the need for a more nuanced understanding of how factors such as aneurysm dimensions and biological sex influence treatment efficacy in aneurysmal populations.

Platelets could play a dual role in the progression of aortic aneurysms, which may be influenced by the size of the aneurysm. In smaller aneurysms, typically less than 50 mm in diameter, the impact of platelets and, by extension, the effect of antiplatelet therapy such as ticagrelor and aspirin, may not result in the attenuation of growth. ^3,24^ This can be attributed to the relatively stable nature of smaller aneurysms, where the pathological processes are less active or extensive. ^1,25^ However, as the aneurysm grows beyond 50 mm, the dynamics change significantly; the increased wall stress and the presence of a larger intraluminal thrombus could provide a more active environment for platelet aggregation and activation. ^2,4,5^ Here, platelets could contribute more actively to inflammatory processes and the remodeling of the extracellular matrix, thus playing a more significant role in aneurysm progression.^1,10,26,27^ This suggests how the effectiveness of antiplatelet therapies would be more evident in larger aneurysms, where the modulation of platelet activity slows the rate of aneurysm expansion and reduce the risk of rupture. ^15^

Our subgroup analysis uncovered a significant interaction between aspirin use and sex in the AAA group, with female patients, on aspirin, exhibiting a marked increase in aneurysm progression. Although AAA is more common in male patients, it is well established that female patients experience worse outcomes. Women face a higher risk of rupture, often at smaller aneurysm diameters, and have poorer outcomes following repair compared with men.^5,28,29^ Our findings align with the sexually dimorphic nature of aneurysms, extending this concept to the context of response to drug therapy. This observation highlights the potential for sex-specific differences in platelet reactivity and response to antiplatelet therapy, underscoring the need for further investigation into personalized approaches for managing aortic aneurysms.

One of the key strengths of our study is the use of a novel AI-based data extraction method, which enabled efficient and accurate retrieval of information from medical records, thereby enhancing the reliability and precision of our findings. The NLP techniques have been demonstrated to be effective and reliable for screening large-scale EHRs ^17,18^ and acquiring structured data from unstructured text. These methods hold significant promise for facilitating patient selection in future epidemiological or comparative effectiveness studies, particularly in the context of aortic aneurysms. Clinical aneurysm research heavily relies on extracting aortic diameter measurements from radiology reports and correlating this data with drug histories and biological markers documented in the records. To the best of our knowledge, our study is the first to employ advanced NLP techniques for extracting aortic diameter data from medical imaging reports while simultaneously exploring this topic with greater granularity and depth than previous studies. This innovative approach demonstrates the potential of AI to enhance the analysis of anonymous medical text data, paving the way for more robust and data-driven research in the field.

Our study offers a comprehensive analysis of platelet counts, aspirin therapy uses, and aneurysm progression, with a particular emphasis on the influence of sex on aneurysm progression in both AAA and TAA patients. By including both aneurysm types, we provide a broader perspective on the role of antiplatelet therapy in these conditions. Despite these strengths, several limitations must be acknowledged. First, accurately determining the timing of aneurysm growth remains challenging, as this process is likely irregular. In our study, aneurysm growth was defined based on the first imaging study showing an increase in size, which may not precisely capture the dynamics of growth over time. Additionally, the retrospective design of our study limits the ability to establish causal relationships. While we observed associations between aspirin therapy and aneurysm progression, these findings require validation through prospective, randomized controlled trials. Another limitation was the relatively small number of patients receiving aspirin therapy, particularly in the TAA group, which reduced statistical power and increased the risk of type II errors. Furthermore, we did not account for differences in aspirin dosing regimens, which may influence therapeutic outcomes. Lastly, our study was confined to data from the University of Kentucky Healthcare system, limiting our ability to report on aneurysm related events, mortality, and cardiovascular outcomes occurring outside this system. We lacked access to information on out-of-hospital deaths or events managed by other healthcare providers, which likely resulted in an underestimation of event rates. As a result, we did not perform comparisons using our database, as it may not accurately reflect the true incidence of events due to the exclusion of external cases. These limitations highlight the need for further research to validate and expand upon our findings, particularly in diverse populations and prospective study designs.

In conclusion, our study, utilizing novel NLP methods for the first time in aneurysm research, contributes to the growing understanding of the role of platelets in aortic aneurysm progression and the potential impact of antiplatelet therapy. By focusing on both AAA and TAA, as well as sex-specific effects, our findings shed light on the complexity of aneurysm management. This study underscores the need for further research to develop tailored therapeutic strategies. Future investigations should prioritize prospective study designs, larger and more diverse patient populations, and in-depth analyses of various antiplatelet agents to provide more definitive evidence on the efficacy of these therapies in aortic aneurysm management.

## Data Availability

All data used will be made available from the Corresponding author upon reasonable request.

## Acknowledgments

The authors’ research was supported by the National Heart, Lung, and Blood Institute of the National Institutes of Health R35HL150818 to S.W.W. Additional support was provided by the NIH National Center for Advancing Translational Sciences through grant numbers UL1TR000117 and UL1TR001998. The content is solely the responsibility of the authors and does not necessarily represent the official views of the NIH. The authors wish to thank the members of the Whiteheart lab for their comments on the manuscript and their careful editing of the final version.

## Authors Contribution

S.M. designed the study, performed all experiments, acquired and analyzed the data, and wrote the manuscript. K.H. and K.M. conducted the statistical analysis, supervised the statistical analysis, and contributed to the statistical results interpretation. H.A. and L.D. assisted with data extraction and manual validation. S.T. supervised the clinical aspects of the study and facilitated data acquisition. S.W.W. is the project leader, overseeing the entire project, and provided critical revisions to the manuscript. All authors reviewed and approved the final manuscript.

## SUPPLEMENTAL MATERIAL

### Code Snippet for NLP-based Data Extraction

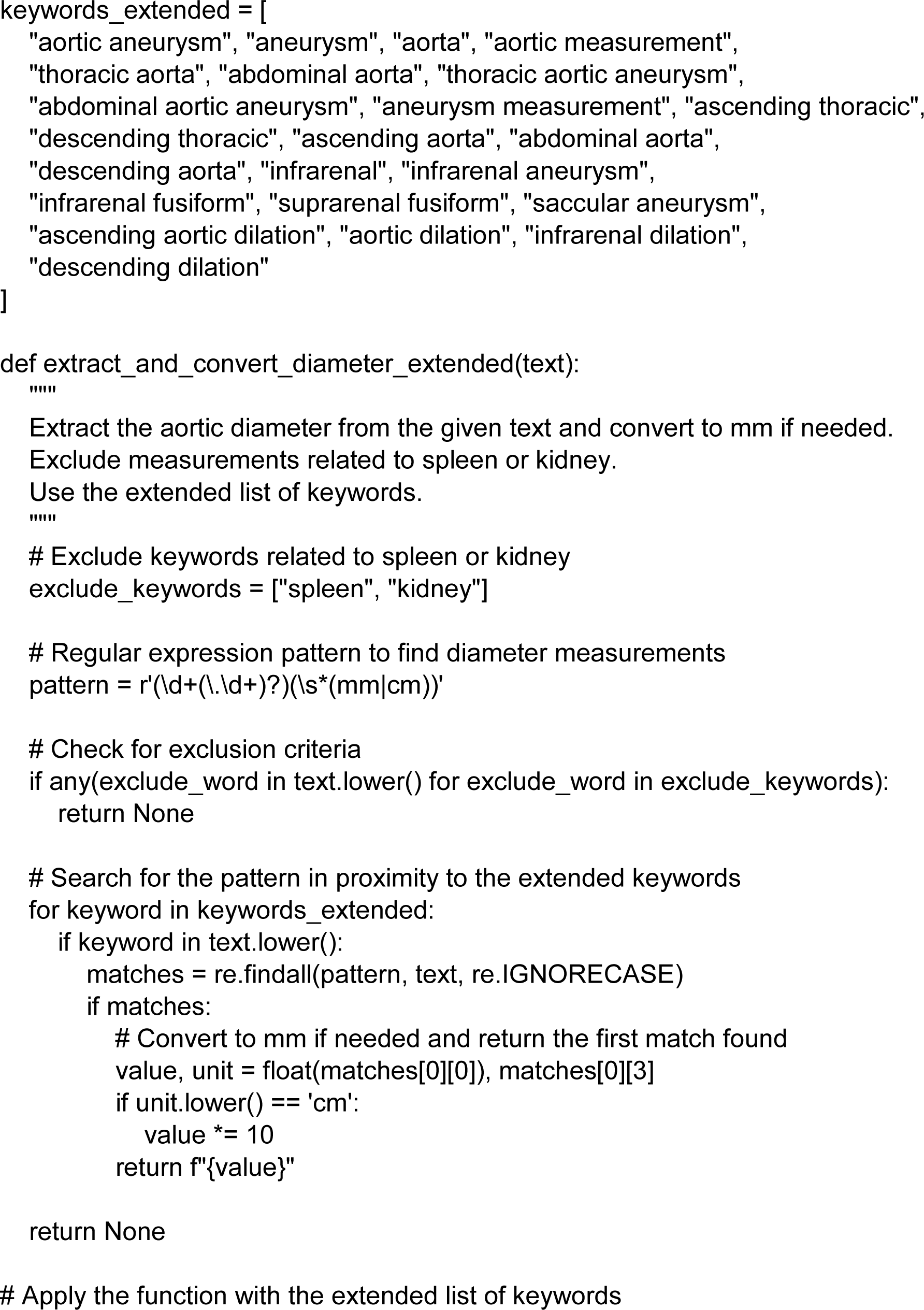
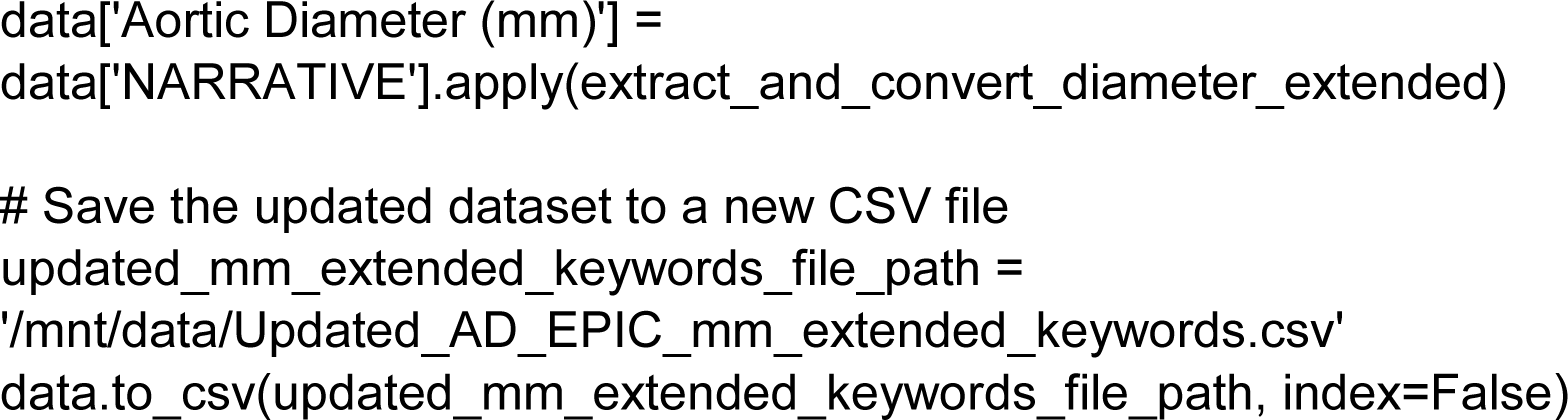

**Supplementary Table 1.** ICD-10 codes for electronic medical records data extraction.

| Variable | Code |
| --- | --- |
| Thoracic aortic aneurysm, ruptured | I71.1 |
| Thoracic aortic aneurysm, ruptured, unspecified | I71.10 |
| Aneurysm of the ascending aorta, ruptured | I71.11 |
| Aneurysm of the aortic arch, ruptured | I71.12 |
| Aneurysm of the descending thoracic aorta, ruptured | I71.13 |
| Thoracic aortic aneurysm, without rupture, unspecified | I71.2 |
| Thoracic aortic aneurysm, without rupture, unspecified | I71.20 |
| Aneurysm of the ascending aorta, without rupture | I71.21 |
| Aneurysm of the aortic arch, without rupture | I71.22 |
| Aneurysm of the descending thoracic aorta, without rupture | I71.23 |
| Abdominal aortic aneurysm, ruptured | I71.3 |
| Abdominal aortic aneurysm, ruptured, unspecified | I71.30 |
| Pararenal abdominal aortic aneurysm, ruptured | I71.31 |
| Juxtarenal abdominal aortic aneurysm, ruptured | I71.32 |
| Infrarenal abdominal aortic aneurysm, ruptured | I71.33 |
| Aortic aneurysm of unspecified site, ruptured | I71.8 |
| Abdominal aortic aneurysm, without rupture | I71.40 |
| Pararenal abdominal aortic aneurysm, without rupture | I71.41 |
| Juxtarenal abdominal aortic aneurysm, without rupture | I71.42 |
| Infrarenal abdominal aortic aneurysm, without rupture | I71.43 |
| Aortic aneurysm of unspecified site, without mention of rupture | I71.9 |

STROBE Statement—Checklist of items that should be included in reports of *cross-sectional studies*
|  | Line Number | Recommendation |
| --- | --- | --- |
| Title and abstract | 1 | (a) Indicate the study’s design with a commonly used term in the title or the abstract |
|  | 31-54 | (b) Provide in the abstract an informative and balanced summary of what was done and what was found |
| Introduction |  |  |
| Background/rationale | 56-74 | Explain the scientific background and rationale for the investigation being reported |
| Objectives | 75-89 | State specific objectives, including any prespecified hypotheses |
| Methods |  |  |
| Study design | 91 | Present key elements of study design early in the paper |
| Setting | 94-112 | Describe the setting, locations, and relevant dates, including periods of recruitment, exposure, follow-up, and data collection |
| Participants | 93-151 | (a) Give the eligibility criteria, and the sources and methods of selection of participants |
| Variables | 93-151 | Clearly define all outcomes, exposures, predictors, potential confounders, and effect modifiers. Give diagnostic criteria, if applicable |
| Data sources/<br>measurement | 114-163 | For each variable of interest, give sources of data and details of methods of assessment (measurement). Describe comparability of assessment methods if there is more than one group |
| Bias | N/A | Describe any efforts to address potential sources of bias |
| Study size | 94-105 | Explain how the study size was arrived at |
| Quantitative variables | 165-190 | Explain how quantitative variables were handled in the analyses. If applicable, describe which groupings were chosen and why |
| Statistical methods | 165-190 | (a) Describe all statistical methods, including those used to control for confounding |
|  |  | (b) Describe any methods used to examine subgroups and interactions |
|  |  | (c) Explain how missing data were addressed |
|  |  | (d) If applicable, describe analytical methods taking account of sampling strategy |
|  |  | (e) Describe any sensitivity analyses |
| Results |  |  |
| Participants | 191-208 | (a) Report numbers of individuals at each stage of study—eg numbers potentially eligible, examined for eligibility, confirmed eligible, included in the study, completing follow-up, and analysed |
|  |  | (b) Give reasons for non-participation at each stage |
|  |  | (c) Consider use of a flow diagram |
| Descriptive data | 209-259 | (a) Give characteristics of study participants (eg demographic, clinical, social) and information on exposures and potential confounders |
|  |  | (b) Indicate number of participants with missing data for each variable of interest |
| Outcome data | 232-255 | Report numbers of outcome events or summary measures |
| Main results | 192-259 | (a) Give unadjusted estimates and, if applicable, confounder-adjusted estimates and their precision (eg, 95% confidence interval). Make clear which confounders were adjusted for and why they were included |
|  |  | (b) Report category boundaries when continuous variables were categorized |
|  |  | (c) If relevant, consider translating estimates of relative risk into absolute risk for a meaningful time period |
| Other analyses | 250-255 | Report other analyses done—eg analyses of subgroups and interactions, and sensitivity analyses |
| **Discussion** |  |  |
| Key results | 261-267 | Summarise key results with reference to study objectives |
| Limitations | 302-316 | Discuss limitations of the study, taking into account sources of potential bias or imprecision. Discuss both direction and magnitude of any potential bias |
| Interpretation | 317-340 | Give a cautious overall interpretation of results considering objectives, limitations, multiplicity of analyses, results from similar studies, and other relevant evidence |
| Generalisability | 341-349 | Discuss the generalisability (external validity) of the study results |
| **Other information** |  |  |
| Funding | 350-357 | Give the source of funding and the role of the funders for the present study and, if applicable, for the original study on which the present article is based |

